# Efficacy and Safety of Iguratimod Combined with Yunke Injection in the Treatment of Ankylosing Spondylitis

**DOI:** 10.64898/2026.03.12.26348262

**Authors:** Zhang Shiyu, Chen Lifeng

**Author notes:** the name of the co-corresponding authors: Zhang Shiyu, Chen Lifeng, e-mail addresses.

## Abstract

**Background:** Biologics and Janus kinase (JAK) inhibitors carry specific risks for Ankylosing Spondylitis patients at risk of tuberculosis infection or those with contraindications such as a history of cancer, there is an urgent need to explore safe and effective alternative treatment options.

**Aims:** To evaluate the efficacy and safety of Iguratimod combined with Yunke injection in the treatment of ankylosing spondylitis at risk of tuberculosis infection or those with a history of cancer.

**Study Design:** Retrospective cohort study.

**Methods:** A retrospective study was conducted on 48 patients with ankylosing spondylitis who had received treatment over the past 3 years and had a history of tuberculosis infection or malignancy. Their treatment regimens and therapeutic outcomes were analyzed, with particular attention to the progression of tuberculosis and malignancy.

**Results:** There was 30 patients receiving Iguratimod combined with Yunke injection treatment, and non-steroidal anti-inflammatory drugs (NSAIDs) were added when pain was severe,referred to as the observation group; 18 patients took Iguratimod and NSAIDs, referred to as the contral group. After treatment of 24 months, both groups showed significant improvements in Ankylosing Spondylitis Disease Activity Score (ASDAS), Bath Ankylosing Spondylitis Functional Index (BASFI), modified Stoke Ankylosing Spondylitis Spine Score (mSASSS), Erythrocyte Sedimentation Rate (ESR), and C-Reactive Protein (CRP), and overall levels could achieve low disease activity. However, the improvement of observation groupin was better than that in the control group, P<0.05. Moreover, the use of NSAIDs in the observation group was significantly less than that in the control group, P<0.001.

**Conclusion:** This study shows that Iguratimod combined with Yunke injection has good efficacy in patients with ankylosing spondylitis who cannot use biologics or JAK inhibitors, not only alleviating pain and morning stiffness but also slowing radiographic progression and reducing the dose of NSAIDs. The combination has a synergistic effect and does not increase adverse reactions. This therapy provides a novel option for patients with specific ankylosing spondylitis.

## 1. Introduction

Ankylosing Spondylitis (AS) is a disease characterized by chronic inflammation of the axial joints, often leading to spinal stiffness, limited mobility, and significant decline in quality of life. Currently, the main treatments for AS include non-steroidal anti-inflammatory drugs (NSAIDs), biologics (such as TNF-α inhibitors and IL-17 inhibitors), and JAK inhibitors [1-3]. However, biologics and JAK inhibitors carry specific risks, including increased susceptibility to active tuberculosis infection, potential tumor risks (especially in patients with a history of cancer), and cardiovascular events (particularly with JAK inhibitors) [4-6]. For patients at risk of tuberculosis infection (e.g., T-SPOT positive) or those with contraindications or concerns such as a history of cancer, there is an urgent need to explore safe and effective alternative treatment options.

Yunke Injection (Technetium [99Tc] Methylenediphosphonate Injection) is a formulation with unique pharmacological properties. Studies demonstrate its multiple effects including immunomodulation, anti-inflammatory action, analgesia, and inhibition of osteoclast activity (with potential anti-osteoporotic effects)[7]. Iguratimoda, small-molecule synthetic drug with immunomodulatory effects, has been shown to exhibit anti-inflammatory, immune-regulating, and bone-protective effects in conditions such as rheumatoid arthritis, with relatively mild immunosuppressive potency[8-9]. The combination of these two agents may synergistically exert anti-inflammatory, immunomodulatory, and bone-protective effects. Current data suggest they have low risks of inducing or exacerbating tuberculosis and tumors, and no significant effects on cardiovascular, cerebrovascular, or herpes zoster conditions.

Against this backdrop, this study aims to evaluate the clinical efficacy and safety of Ailamod combined with Yunke Injection in a specific population of AS patients (those unable or unwilling to use biologics and JAK inhibitors due to risks such as tumors or tuberculosis infection) through a retrospective analysis.

## 2. Materials and Methods

### 2.1 Study Subjects

#### 2.1.1 Study Design

A single-center retrospective cohort study

#### 2.1.2 Case Sources

AS patients treated in the outpatient or inpatient department of authors’ hospital between January 1, 2020 and December 31,2023. Data sources included admission records, medical course records, discharge summaries, outpatient records, laboratory reports, imaging reports, nursing records, and telephone follow-up records.

#### 2.1.3 Inclusion Criteria

(1) Compliance with the 1984 revised New York Classification of Ankylosing Spondylitis. (2) Age range: 18-70 years. (3) Disease in active phase: Ankylosing Spondylitis Disease Activity Score (ASDAS)≥2.1. (4) Treatment regimen: Either: (a) Iguratimod combined with Yunke Injection for≥24 months; or (b) Iguratimod combined with on-demand NSAIDs for≥24 months. (5) No prior use of methotrexate, sulfasalazine, thalidomide, Tripterygium wilfordii preparations, biologics, JAK inhibitors, or glucocorticoids within 6 months before study initiation, with only NSAIDs for anti-inflammatory and analgesic purposes.

#### 2.1.4 Exclusion Criteria

(1) Failure to meet any of the above inclusion criteria. Non-axis type, non-radiating. (2) Concurrent severe hepatic, renal, or cardiac insufficiency (such as Child-Pugh C grade cirrhosis, ALT elevated by 2 times, eGFR <45 ml/min/1.73m², cardiac function grade 4) or hematological disorders (such as severe anemia, agranulocytosis, thrombocytopenia, etc.). (3) Pregnant or breastfeeding women. (4) Concurrent use of immunomodulatory traditional Chinese medicine preparations during the study period.

#### 2.1.5 Sample Screening

A total of 1,800 hospitalized AS patients from 2020 to 2023 were screened. Following the aforementioned stringent inclusion and exclusion criteria, 30 eligible patients receiving combination therapy were selected as the observation group for analysis. Additionally, 18 AS patients receiving only Iguratimod and NSAIDs were identified as the control group. Authors hadn’t access to information that could identify individual participants during or after data collection.

The observation group comprised 24 males and 6 females aged 20-55 years, with disease duration ranging from 0.5 to 10 years. Among them, 15 patients had latent tuberculosis with T-spot reactivity during treatment and a history of pulmonary tuberculosis, while 5 were thyroid cancer resected patients and 2 were lung cancer resected patients. The control group included 15 males and 3 females aged 21-56 years, with disease duration of 0.5 to 8 years. Six patients had latent tuberculosis with T-spot reactivity during treatment and a history of pulmonary tuberculosis, and 1 was a thyroid cancer resected patient. No significant differences were observed in age or disease duration between the two groups (p>0.05), as shown in Table 1.

**Table 1.** Summary of baseline data.

| index | observation group<br>(n=30) | control group (n=18) | P value |
| --- | --- | --- | --- |
| sex | 24 males and 6 females<br>(male: female = 4:1) | 15 males, 3 females<br>(male: female = 5:1) |  |
| age | 29.2±8.8 years<br>IQR: 27.5 (25-31) | 29.5±9.3 years<br>IQR: 27 (25-31) | 0.85 |
| course of disease | 2.40±2.04 years<br>IQR: 2.45 (1.7-3.2) | 2.35±1.69 years<br>IQR: 2.35 (1.5-3.0) | 0.92 |
| mSASSS, mean value<br>(SD) | 16.43 ± 6.96<br>IQR: 15.5 [8, 32] | 15.56 ± 6.44<br>IQR: 13.5[1, 24] | t = 0.470,<br>p = 0.641 |
| ligamentous<br>osteophytes present,<br>n (%) | 19 (63.3%) | 11 (61.1%) | p = 0.878 |

### 2.2 Treatment Protocol

2.2.1 All patients in the observation group received the following combination therapy: (1) Iguratimod (China Hainan Xiansheng Pharmaceutical Co., Ltd. production): Oral administration at 25mg twice daily.

(2) Yunk Injection (Technetium [99Tc] Methylenediphosphonate Injection, Chengdu Yunke Pharmaceutical Co., Ltd. production): Intravenous infusion in once daily doses (each dose containing 0.15μg [99Tc], 15mg methylenediphosphonic acid, and 1.5mg stannous chloride), dissolved in 250ml 0.9% sodium chloride injection. One dose per day.

(3) Concurrent Medication: NSAIDs may be administered as needed when pain becomes intolerable to relieve symptoms.

2.2.2 The 18 patients in the control group received Iguratimod and NSAIDs as needed under the same protocol as the observation group.

2.2.3 The equivalence conversion of NSAIDs used in this study is as follows: imrecoxib (0.1g, bid) = Celecoxib (0.2g, bid) = Loxoprofen Sodium (60mg, bid) = Etoricoxib (120mg, qd).

### 2.3 Observation Indicators

#### 2.3.1 Baseline Data

1. Demographic data: Age, sex, disease duration.
2. Disease characteristics: Disease activity score calculated as ASDAS-CRP =0.12 × back pain score + 0.06 × morning stiffness duration + 0.11 × overall patient score + 0.07 × peripheral joint score + 0.58 × ln(CRP + 1), score <1.3 indicates remission, 1.3-2.1 indicates low activity, 2.1-3.5 indicates high activity, and>3.5 indicates extremely high activity. (ln = natural logarithm; CRP unit: mg/L). Functional index (BASFI): 0-10 points (higher score indicates more severe functional impairment). Inflammatory markers: CRP (mg/L), ESR (mm/h). Imaging assessment: The mSASSS score is applied to spinal X-rays (lateral views of cervical and lumbar vertebrae). Two experienced radiologists independently evaluate the images, with the average score calculated. The assessment covers 8 anterior vertebral margins and 16 scoring points, where 0 indicates no injury and 48 indicates complete spinal ankylosis. Scoring criteria: <10 points: mild structural injury; 10-30 points: moderate injury;>30 points: severe injury. Complications: Specifically, factors that may prevent the use of biologics/JAK inhibitors (e.g., uncontrolled tuberculosis infection or a history of high-risk tumors within 5 years).
3. Baseline laboratory safety indicators: Complete Blood Count (CBC): White blood cell count (WBC),Neutrophil count (NEUT),Ly mphocyte count (LYM),Hemoglobin (HGB),Platelet count (PLT). Liver function tests: Alanine aminotransferase(ALT),Aspartate aminotransferase (AST), Total bilirubin (TBIL),Albumin (ALB). Renal function tests: Serum creatinine (Scr), Estimated glomerular filtration rate (eGFR), Blood urea nitrogen (BUN). Urine analysis. Tuberculosis screening results (e.g., T-SPOT, PPD test, chest CT report). Cancer biomarkers (selected based on patient’s tumor history or high-risk factors): CEA (carcinoembryonic antigen),AFP (alpha-fetoprotein),CA19-9 (carcinoembryonic antigen 19),PSA (prostate-specific antigen).

#### 2.3.2 Efficacy evaluation metrics (assessment time points): T0 (baseline),T24 (24 months)

1. Primary efficacy measures: ASDAS-CRP changes: Calculate the change in T24 relative to T0. Evaluate the proportion of patients achieving ASDAS clinically important improvement (CII: decrease≥1.1), major improvement (MI: decrease≥2.0), low disease activity (LDA: ASDAS <2.1), and inactive disease (ID: ASDAS <1.3).
2. Secondary efficacy measures: BASFI changes: Calculate the change in T24 relative to T0. Inflammatory marker changes: Calculate the change in CRP (mg/L) and ESR (mm/h) relative to T0 at T24, along with normalization rates.
3. NSAID usage: Total NSAID dosage over 24 months (converted to celecoxib equivalent dose).
4. Imaging progression: Progression is defined as an increase of≥2 points in the mSASSS score. Calculate the proportion of patients with newly developed osteophytes or ligament ossification at the anterior spinal margin.

#### 2.3.3 Safety measures (during 24-month treatment period)

1. Adverse Events (AEs): Document all reported or recorded adverse events, regardless of their potential drug association. Detailed records must include: the time of occurrence, severity (mild, moderate, or severe), duration, specific manifestations/symptoms, measures taken (e.g., symptomatic treatment, dose reduction, temporary discontinuation, or permanent withdrawal), and outcomes (remission, no remission, or sequelae).
2. Adverse Events of Special Interest (AESIs): Infections: Incidence and severity of non-cluster upper respiratory tract infections, urinary tract infections, and gastrointestinal infections. COVID-19 pandemic excluded, as all individuals were infected with SARS-CoV-2. Tuberculosis infection/activation: Cases of newly active tuberculosis or reactivated latent tuberculosis (symptoms, diagnostic criteria including sputum smear/culture/X-ray/CT/PCR, treatment, and outcomes). Follow-up T-SPOT/TB-ELISPOT results. Shingles: Location, severity, treatment, and outcomes. Other severe/occlusive infections. Hematological toxicity: Incidence, severity, and management of leukopenia (<3.5×10^9^/L), neutropenia (<1.8×10^9^/L), anemia (HGB decrease ≥20g/L or below normal value), and thrombocytopenia (<100×10^9^/L). Hepatic dysfunction: Incidence, severity, management (whether to discontinue the drug), and recovery status of ALT/AST elevation (>1x ULN,>2x ULN,>3x ULN). Record the elevation of total bilirubin. Renal dysfunction: Incidence and management of Scr elevation (>1.5x baseline value or>1.5x ULN) and eGFR reduction (>20% decrease from baseline or <60 ml/min/1.73m²). Gastrointestinal reactions: Incidence and severity of nausea, vomiting, abdominal pain, diarrhea, and dyspepsia. Skin reactions: Incidence and severity of rash and pruritus. Cardiovascular events: Incidence of new or worsening hypertension, myocardial infarction, stroke, heart failure, deep vein thrombosis/pulmonary embolism, etc. (with detailed documentation of diagnosis, treatment, and outcomes). Malignant tumors: New malignant tumors or recurrence/metastasis of existing tumors (including tumor type, diagnosis date, stage, and treatment).
3. Drug tolerance: Proportion and reasons for dose reduction due to adverse events. Proportion, reasons, and duration of temporary discontinuation due to adverse events. Proportion and reasons for permanent discontinuation due to adverse events.
4. Serious Adverse Events (SAEs): All events causing death, life-threatening conditions, hospitalization or prolonged hospital stays, persistent or significant functional impairment, or congenital malformations/birth defects.

#### 2.3.4 Treatment Compliance

Record the actual number of annual treatment courses completed for Yunke Injection in the observation group (whether all three small cycles were completed each year). Document discontinuation of Iguratimod in both groups (reasons and duration). Assess overall medication adherence (good/average/poor) through prescription records and medical records.

#### 2.3.5 Ethics

The retrospective study protocol was approved by the ethics committee, with patient informed consent waived.

### 2.4 Statistical Methods

#### 2.4.1 Data Preprocessing

(1) Missing Value Handling: Cases with over 20% missing values in key variables (e.g., ASDAS-CRP, radiological examinations) were excluded. Secondary variables with missing values were imputed using multiple imputation. (2) Outlier Detection: Identifying outliers through box plots or standard deviation methods (e.g.,±3SD), with clinical judgment applied to determine retention or correction.(3)Data Transformation: Non-normally distributed continuous variables (e.g., CRP, ESR) were logarithmically transformed to meet parameter testing requirements.

#### 2.4.2 Baseline Data Analysis

(1) Descriptive Statistics: For continuous variables: Mean±Standard Deviation (Mean±SD) for normally distributed data, median (Interquartile Range, IQR) for non-normally distributed data. Categorical variables: Frequency and percentage (n,%). (2) Between-group comparability test: Compare observation group (combined therapy) vs control group (almodriptan monotherapy). For continuous variables: Independent samples t-test for normally distributed and homoscedastic data; Mann-Whitney U test for non-normally distributed or non-homoscedastic data. For categorical variables:χ²test or Fisher’s exact test (when expected frequency <5).

#### 2.4.3 Primary efficacy analysis (Key focus: Time effect & group differences)

Primary endpoint: Changes in ASDAS-CRP and BASFI scores at baseline (T0) and 24 months (T24).

Statistical analysis method:

1. Within-group comparison (pre-and post-treatment changes): Paired t-test (for normal data) or Wilcoxon signed-rank test (for non-normal data) to compare T24 vs T0. Calculate effect size: Cohen’s d (t-test) or r = Z/√N (Wilcoxon) to quantify change magnitude.
2. Between-group comparison (combination vs monotherapy): Linear Mixed Model (LMM). Dependent variable: ASDAS or BASDAI. Fixed effects: Group (combination/monotherapy), time point (T0/T24), group × time interaction term (if significant, indicating different time trends between groups). Random effect: Patient ID (considering intra-individual measurement correlation). Adjusted variables: Baseline covariates (e.g., age, disease duration, baseline ASDAS). Alternative approach (for small samples or complex models): Calculate change values (Δ) from T0 to T24 for each group, then compare Δ differences between groups using independent samples t-test.

#### 2.4.4 Secondary Outcomes Analysis

1. Continuous Variables (CRP, ESR, etc.): Methods are identical to primary outcomes (logistic regression model [LMM] or between-group comparisons of change values).
2. Categorical Response Rates (e.g., ASDAS-CII/MI/LDA/ID rates, NSAID discontinuation rates): Within-group comparisons: McNemar test (for paired categorical data, e.g., T24 vs T0 response rates). Between-group comparisons: Fisher’s exact test (direct comparison of response rates at T24).
3. Imaging Progression (mSASSS): Within-group comparisons: paired t-test. Between-group comparisons: independent t-test. Progression rate (ΔmSASSS ≥ 2): Fisher’s exact test.

#### 2.4.5 Safety Analysis

1. Adverse Event (AE) Description: Calculate incidence rates by categorizing events by type, severity, and drug-relatedness. For common AEs (e.g., infections, elevated liver enzymes), perform Fisher’s exact test to compare incidence rates between groups.
2. Laboratory Parameter Abnormalities: Analyze the proportion of abnormal values at each time point (e.g., ALT>1×ULN ratio). Use linear mixed models (LMM) or repeated measures ANOVA to compare time-dependent changes between groups.
3. Serious Adverse Events (SAEs) and Of Special Concern Adverse Events (AESIs): Provide detailed case descriptions, calculate incidence rates, and interpret cautiously (rare events may not be detected in small samples).
4. Drug Tolerance (Dose Reduction/Discontinuation Rate): Compare groups using Fisher’s exact test.

#### 2.4.6 Statistical Software and Significance Criteria

The study employed SPSS 26.0 (IBM, USA) with a two-tailed significance level of α = 0.05. Bonferroni correction (α = 0.025) was applied for multiple comparisons in the primary endpoint (ASDAS/BASDAI at T24).

## 3. Results

The observation group demonstrated superior improvements in inflammatory markers, ASDAS, BASFI, and mSASSS compared to the control group.

### 3.1 Inflammatory Markers Level

Before treatment, no significant difference was observed in the mean values of CRP and ESR between the two groups. After treatment, intra-group comparisons showed a significant decrease in both CRP and ESR levels (P<0.01). Inter-group comparisons revealed that the mean CRP level in the observation group was significantly lower than that in the control group (6.29±3.02 vs. 13.92±6.87, t=4.52, P<0.001, Cohen’s d=1.67), while the mean ESR level in the observation group was significantly lower (16.07±5.68 vs. 20.72±6.55, t=−2.57, p=0.013, Cohen’s d=0.75). These results indicate that the observation group exhibited significantly lower inflammatory markers than the control group after treatment.

### 3.2 AS Disease Activity

Before treatment, the mean ASDAS scores in the observation group were 3.02±0.68 compared to 2.89±0.59 in the control group, with no significant baseline difference (t=0.72, P=0.48), indicating balanced group distribution. After treatment, ASDAS scores decreased to 1.62±0.31 in the observation group and 1.72±0.26 in the control group. Both groups showed significant post-treatment score reductions (observation group: t=11.72, P<0.001; control group: t=9.85, P<0.001), demonstrating the intervention’s effectiveness. However, independent samples t-test revealed no statistically significant difference in post-treatment scores between groups (t=−1.32, df=46, P=0.19), with a mean difference of-0.10 (95% CI: −0.25 to 0.05). Although the observation group’s mean was slightly lower, the small effect size (Cohen’s d=0.34) suggests limited clinical significance. Supplementary sensitivity analysis using Welch’s t-test (for heteroscedasticity) confirmed these findings (t=−1.44, df=30.7, P=0.16).

### 3.3 AS Function Index

Before treatment, the mean BASFI scores in the observation group were 2.83±0.61 compared to 2.89±0.59 in the control group, with no significant baseline difference (t = −0.35, P=0.73), indicating balanced group distribution. After treatment, the BASFI scores in the observation group decreased to 1.69±0.20 and 2.10±0.26 in the control group, respectively. Both groups showed significant post-treatment score reductions from baseline (observation group: t=11.72, P<0.001; control group: t=9.75, P<0.001), demonstrating the intervention’s effectiveness in both groups. The independent samples t-test was used to compare the differences between the two groups after treatment. After Welch’s correction, the difference between the two groups was statistically significant (t=−6.982, df=28.74, P<0.001).

### 3.4 Imaging changes

The pre-treatment spinal mSASSS scores were essentially identical in both groups, as shown in Table 1. No significant difference was observed in baseline mSASSS levels between the two groups.

After treatment,the observation group demonstrated a statistically significant reduction in mSASSS scores (P<0.001) with a large effect size (d=2.41), while the control group also showed marked improvement (P<0.001). However, the reduction in the observation group was significantly greater than that in the control group (mean difference Δ=0.31,95% CI: 0.12–0.50, P=0.002). Post-treatment intergroup comparison revealed a Δ=0.36 (95% CI [0.22,0.50]), t=4.32, P<0.001, with a Cohen’s d=1.52 (95%CI [0.89,2.15]), supporting the observation group’s superior efficacy. Covariance analysis (ANCOVA) confirmed the significant intergroup difference even after adjusting for baseline mSASSS (F=15.63, P<0.001).As shown in Table 2.

**Table 2.**
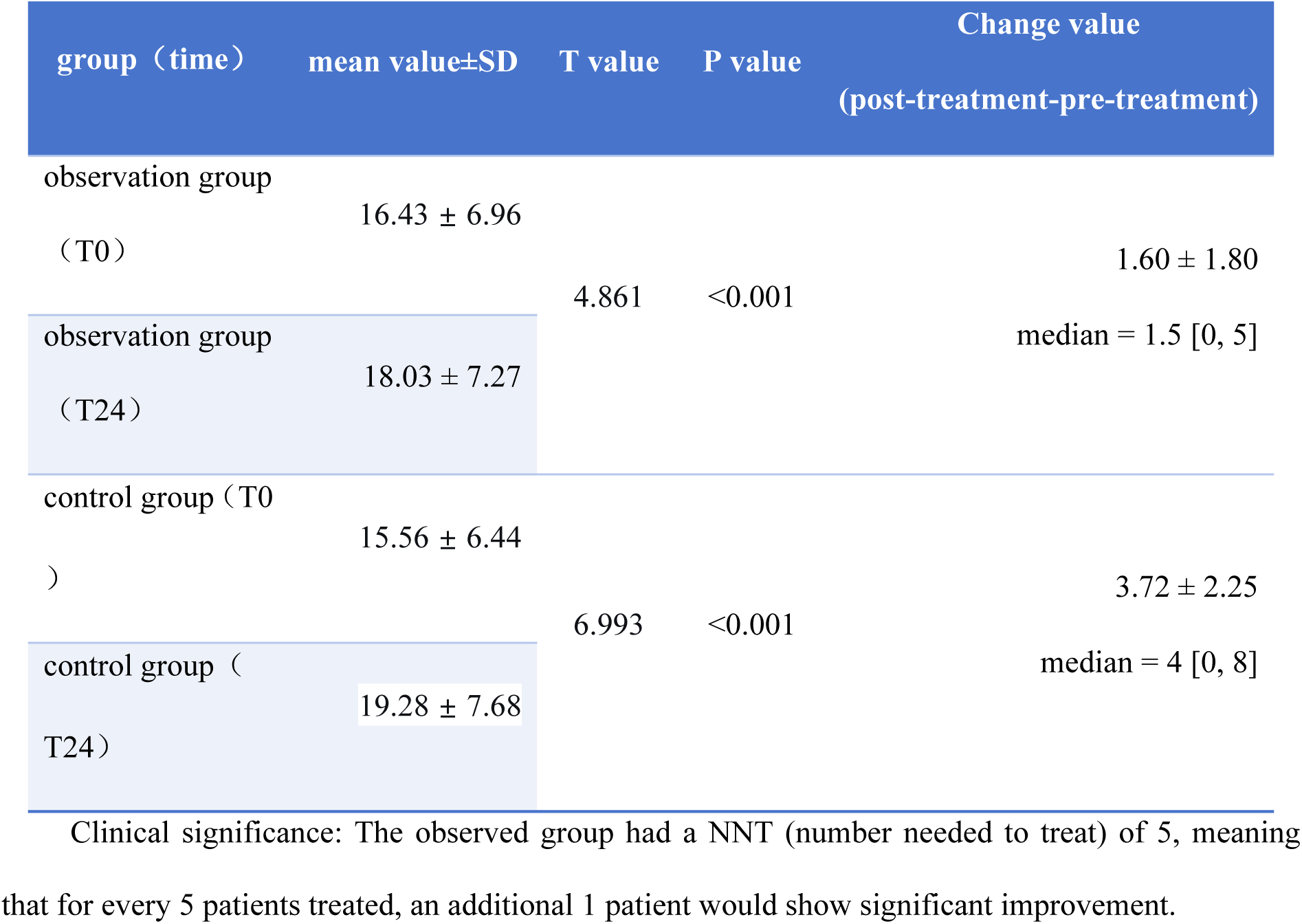
Comparison of mSASSS scores before and after treatment in two groups.

### 3.5 NSAID dosage after treatment

**Table 3.** Comparison of NSAID dosage (g) between the two groups after 24 months.

| group | mean value±SD<br>(g) | 95%CI | T value | P value | Cohen's d<br>(95%CI) |
| --- | --- | --- | --- | --- | --- |
| observation group (T24) | 128.6±27.3 | [118.4, 138.8] | 9.27 | <0.001 | 2.83 (1.98–3.68) |
| control group (T24) | 210.7±31.5 | [195.2, 226.2] |  |  |  |

Note: The dosage of NSAIDs in the observation group was significantly lower than that in the control group (mean difference 82.1g), and the dispersion of the data in the observation group was
smaller (lower SD).

### 3.6 Adverse event

In the observation group, two patients exhibited elevated transaminase levels (not exceeding three times the upper limit). These patients were concurrently taking atorvastatin and aspirin and had fatty liver disease. The cause of the elevated transaminase levels could not be determined. After two weeks of treatment with silybin capsules, transaminase levels return to normal, while the iguratimod not discontinued or reduced. No cases of tuberculosis recurrence or exacerbation, tumor recurrence, metastasis, or new onset were observed.

In the control group, two patients fused fatty liver developed elevated transaminase levels (within 3-fold upper limit). After two weeks of silybin capsule treatment, while iguratimod was adjusted to one daily tablet for two weeks, restoring normal transaminase levels. Iguratimod was then resumed at standard dosage without adverse reactions.

No severe opportunistic infections, tuberculosis infection/relapse, herpes zoster, hematologic toxicity, renal impairment, gastrointestinal reactions, cardiovascular events, or malignancies occurred in either group. The medications were not discontinued or reduced.

## 4. Discussion and Conclusion

The screening process of 30 patients in this observation group (representing 1.67% of AS patients during the same period) highlighted the specific patient population eligible for this regimen (excluding those who refused biologic agents or JAK inhibitors). Analysis of the combined use of iguratimod and Yunke Injection during 24-month periodic long-term treatment demonstrated significant improvements in AS disease activity (ASDAS, CRP, ESR), functional capacity (BASFI), spinal osteophytes, and NSAID dosage. The results showed comparable efficacy to literature reports on biologic agents and JAK inhibitors [10-13].

In its low oxidation state, the trace element technetium-99m readily gains and loses electrons to neutralize free radicals in the human body. This process maintains the activity of superoxide dismutase (SOD), inhibits the formation of pathological complexes, and prevents tissue damage caused by free radicals. Additionally, when combined with methylenediphosphate, technetium exhibits immunosuppressive effects, demonstrating the properties of a slow-acting anti-rheumatic drug. Methylenediphosphate possesses non-steroidal anti-inflammatory and adrenocortical hormone-like effects, exerting potent anti-inflammatory and analgesic actions by suppressing prostaglandin production and histamine release. It also exhibits tetracycline-like effects: through binding with divalent metal ions, it inhibits collagenase activity in connective tissues, preventing collagen degradation in articular soft tissues. Furthermore, it significantly suppresses osteoclast activity, inhibits bone resorption, repairs cartilage damage caused by various types of arthritis, restores joint function, and promotes tissue repair and regeneration [14,15].

As a novel immunosuppressant, iguratimod exerts anti-inflammatory effects by inhibiting inflammatory cell proliferation and reducing cytokine release, thereby modulating anti-inflammatory signaling pathways. It regulates immune function by influencing immune cell proliferation and cytokine expression, thereby decreasing immune complex production and deposition. Additionally, it exerts bone-protective effects by regulating bone metabolism through Wnt/β-catenin, Toll-like receptor 4/nuclear factor κB, and osteoprotegerin/nuclear factor κB receptor activator ligand signaling pathways. The efficacy and safety of iguratimod have been validated in clinical applications for rheumatoid arthritis and primary Sjögren’s syndrome, and it has been incorporated into disease management guidelines [8-9].

Pharmacological analysis of iguratimod and Yunke Injection reveals their synergistic effects. This combination not only relieves pain but also delays disease progression, reduces NSAID usage, and avoids adverse reactions. The regimen demonstrates favorable safety profiles, particularly showing no adverse effects on tuberculosis reactivation, cancer risk, or infections (e.g., shingles). It holds potential safety advantages for high-risk populations with cancer or tuberculosis.

The limitations of this study include retrospective design, small sample size, and potential selection bias. Future studies should adopt prospective randomized controlled trials with larger sample sizes and longer follow-up periods.

## Data Availability

All relevant data are within the manuscript and its Supporting Information files.

## Statement

All authors have no conflict of interest.

## references

[1] Zhao SS, Harrison SR, Thompson B, et al. The 2025 British Society for Rheumatology guideline for the treatment of axial spondyloarthritis with biologic and targeted synthetic DMARDs. Rheumatology (Oxford). 2025 Jun 1;64(6):3242–3254. doi: 10.1093/rheumatology/keaf089.

[2] Bautista-Molano W, Fernández-Ávila DG, Brance ML, et al. Pan American League of Associations for Rheumatology recommendations for the management of axial spondyloarthritis. N at Rev Rheumatol. 2023 Nov;19(11):724–737. doi: 10.1038/s41584-023-01034-z. Epub 2023 Oct 6. PMID: 37803079.

[3] Rohekar S, Pardo JP, Mirza R, et al. Canadian Rheumatology Association/Spondyloarthritis Research Consortium of Canada Living Treatment Recommendations for the Management of Axial Spondyloarthritis. J Rheumatol. 2025 Jan 1;52(1):10–22. doi: 10.3899/jrheum.2023-1237. PMID: 38950949.

[4] Kwon OC, Lee HS, Jeon SY, et al. Effect of TNF inhibitors on the risk of cancer recurrence in patients with ankylosing spondylitis: a nested case-control study. Rheumatology (Oxford). 2025 Jun 6:keaf313. doi: 10.1093/rheumatology/keaf313.

[5] He W, Yang H, Yang X, et al. Global research trends in biological therapy for ankylosing spondylitis: A comprehensive visualization and bibliometric study (2004-2023). Hum Vaccin Immunother. 2025 Dec;21(1):2445900. doi: 10.1080/21645515.2024.2445900. Epub 2025 Jan 15.

[6] Picchianti-Diamanti A, Aiello A, De Lorenzo C, et al. Management of tuberculosis risk, screening and preventive therapy in patients with chronic autoimmune arthritis undergoing biotechnological and targeted immunosuppressive agents. Front Immunol. 2025 Feb 3;16:1494283. doi: 10.3389/fimmu.2025.1494283.

[7] Wang H, Wu Q, Zhao S. Efficacy Observation of Yunke Injection in the Treatment of Rheumatoid Arthritis (Attached with Report of 36 Cases) [J]. Journal of Jilin University (Medical Edition),2009,35(05):856.

[8] Zeng L, He Q, Deng Y, et al. Efficacy and safety of iguratimod in the treatment of rheumatic and autoimmune diseases: a meta-analysis and systematic review of 84 randomized controlled trials. Front Pharmacol. 2023 Dec 7;14:1189142.

[9] Long Z, Zeng L, Yang K, et al. A systematic review and meta-analysis of the efficacy and safety of iguratimod in the treatment of inflammatory arthritis and degenerative arthritis. Front Pharmacol. 2024 Oct 10;15:1440584.

[10] Tichý Š, Nekvindová L, Baranová J, et al. Drug survival analysis of etanercept compared with monoclonal antibody tumour necrosis factor-α inhibitors in rheumatoid arthritis, psoriatic arthritis, and ankylosing spondylitis: a propensity score-matched analysis from the Czech ATTRA registry. Scand J Rheumatol. 2025 Mar;54(2):79–86. doi: 10.1080/03009742.2024.2381746.

[11] Zemrani S, Amine B, El Binoune I, Rostom S, Tahiri L, Allali F, Bahiri R. The Retention Rate and Safety of Secukinumab as a First-Line Biologic Agent in Axial Spondyloarthritis Compared to a First Tumor Necrosis Factor (TNF) Inhibitor: A Real-World, Longitudinal Study. Cureus. 2024 Sep 28;16(9):e70365.doi: 10.7759/cureus.70365.

[12] Lopalco G, D’Antonio A, Chimenti MS, et al. Upadacitinib for the treatment of radiographic axial spondyloarthritis - case series and review of the literature. Drugs Context. 2025 May 12;14:2024–12-3. doi: 10.7573/dic.2024-12-3.

[13] Baraliakos X, van der Heijde D, Sieper J, et al. Efficacy and safety of upadacitinib in patients with active ankylosing spondylitis refractory to biologic therapy: 2-year clinical and radiographic results from the open-label extension of the SELECT-AXIS 2 study. Arthritis Res Ther. 2024 Nov 12;26(1):197. doi: 10.1186/s13075-024-03412-8.

[14] Long Z, Deng Y, He Q, et al. Efficacy and safety of Iguratimod in the treatment of Ankylosing Spondylitis: A systematic review and meta-analysis of randomized controlled trials. Front Immunol. 2023 Mar 3;14:993860. doi: 10.3389/fimmu.2023.993860. PMID: 36936924; PMCID: PMC10020631.

[15] Liu S, Cui Y, Zhang X. Molecular mechanisms and clinical studies of iguratimod for the treatment of ankylosing spondylitis. Clin Rheumatol. 2021 Jan;40(1):25–32. doi: 10.1007/s10067-020-05207-z. Epub 2020 Jun 6. PMID: 32506313.

